# KNOWLEDGE, ATTITUDES AND PRACTICES OF REFUSE DISPOSAL IN OKELELE, ILORIN, NIGERIA

**DOI:** 10.64898/2026.11.04.26367307

**Authors:** Ibrahim Muhammad, Abdulrahman Salahudeen Alatare, Mariam Oyinkansola Adewunmi, Abdulmalik Opeyemi Adeyemo, David-Okoro Joshua Precious, Maryam Omowumi Adegoke, Asiat Oluwafisayo Adeleke, Idayat Omotoyosi Akindeji

**Author notes:** **Corresponding author** Abdulmalik, Opeyemi, Adeyemo, Department of Medicine, College of Health Sciences, University of Ilorin, P.M.B. 1515, Ilorin, Nigeria. **Platforms For Sharing Your Work Professionally And Socially** LinkedIn: Abdulmalik Adeyemo Twitter: Abdulmalik Adeyemo FaceBook: Abdulmalik Adeyemo ResearchGate: Abdulmalik Adeyemo.

## Abstract

**BACKGROUND AND OBJECTIVE:** Improper refuse disposal remains an environmental health concern in many Nigerian communities. This study assessed knowledge, attitudes and practices relating to refuse disposal among adult residents of Okelele community, Ilorin East Local Government Area, Kwara State, Nigeria.

**METHODS:** A descriptive cross-sectional community-based study was conducted among 180 adults. Data were collected using a structured questionnaire covering sociodemographic characteristics, knowledge, attitudes and disposal practices. Descriptive statistics summarised responses, and chi-square tests assessed associations between categorical variables at a 5% significance level.

**RESULTS:** Of 180 respondents, 102 (56.7%) had good knowledge, while 87 (48.3%) had negative attitudes and 99 (55.0%) had poor refuse-disposal practices. Commonly reported practices included incineration (50.6%), disposal into drains (30.6%) and open burning (21.7%). Absence of waste containers (37.2%) and distance to available containers (28.9%) were the leading reported barriers. Knowledge was associated with practice (χ^2^=22.122; p<0.001) and attitude (χ^2^=8.221; p=0.016); practice was also associated with attitude (χ^2^=5.970; p=0.015).

**CONCLUSION AND IMPLICATIONS FOR TRANSLATION:** Despite generally good knowledge, poor disposal practices were common. Improving access to reliable waste-collection infrastructure, alongside community education and enforcement of local waste-management measures, may help translate knowledge into safer practices.

## 1. INTRODUCTION

Inadequate municipal solid-waste management is an important environmental health challenge, particularly where population growth and urbanisation outpace the provision of collection, transport and disposal services. Uncollected or improperly discarded refuse can accumulate in streets and drainage channels, contribute to local pollution and flooding, and create conditions that support disease vectors.^[1]^ The scale and consequences of these problems depend not only on residents’ knowledge and behaviour but also on the availability and accessibility of waste-management services.^[2,3,4]^

In Nigeria, households use a range of refuse-disposal methods, including collection in designated containers, recycling, incineration, open dumping, burning and disposal into drains. Some practices may expose residents and the wider environment to avoidable risks.^[2]^ Community-level studies have documented gaps between awareness of appropriate waste management and the disposal options that residents can realistically access. Research in Ilorin has also described spatial and service-related challenges in municipal solid-waste management.^[4,5]^

Okelele is a residential community in Ilorin East Local Government Area, Kwara State. The project underlying this article identified inadequate access to public refuse containers and indiscriminate disposal as concerns in the community. Local evidence on residents’ knowledge, attitudes and practices can help inform targeted environmental-health interventions. This study therefore assessed knowledge of refuse disposal, attitudes towards proper disposal, disposal practices and perceived barriers among adult residents of Okelele community.^[6]^

## 2. METHODS

### 2.1 Study design and setting

A descriptive cross-sectional community-based study was conducted in Okelele, Ilorin East Local Government Area, Kwara State, Nigeria. Okelele was described in the source project as a predominantly residential community with common livelihoods including pottery, hand weaving and petty trading, and without an official refuse-dump site.

### 2.2 Study population and eligibility

The study population comprised adults aged 18 years or older who were resident in Okelele. Eligibility required residence in the community for at least two years and willingness to provide informed consent. Individuals younger than 18 years and those unable to participate or unwilling to consent were excluded.

### 2.3 Sample size and sampling

The project reported a calculated minimum sample size of approximately 180 using the formula n=Z^2^pq/d^2^, with Z=1.96, p=0.135, q=0.865 and d=0.05. It also reported an inflation to 200 to allow for non-response, while the final achieved sample was 180. A three-stage sampling procedure was described: Okelele was divided into five zones; houses were selected systematically; households were selected by simple random selection; and one eligible adult was selected in each participating household. The final zone allocation was 30 in Lowin, 37 in Amuyankan, 33 in Omoboriowo, 41 in Jagun and 39 in Babaladifa.

### 2.4 Data collection instrument and procedure

Data were collected using a structured, semi-open-ended questionnaire with four sections: sociodemographic characteristics, knowledge of refuse disposal, attitudes towards refuse disposal, and disposal practices. The questionnaire was reviewed by a professional in the Department of Epidemiology and Community Health, University of Ilorin Teaching Hospital, for face and content validity. It was pretested among 18 adults in Agbabiaka community, Ilorin South Local Government Area, and revised in response to feedback. The project spanned for eight weeks between August and October, with questionnaires administered on Saturdays and Sundays only.

### 2.5 Study variables and statistical analysis

The principal outcomes were respondents’ classified knowledge, attitude and practice levels. Other variables included sociodemographic characteristics, reported disposal methods, perceived consequences of improper disposal and barriers to appropriate disposal. Categorical data were summarised using frequencies and percentages. Associations between knowledge, attitude and practice categories were examined using chi-square tests, with statistical significance set at p<0.05. The statistical software used for the project was Statistical Package for Social Science (SPSS) version 26 and Microsoft Excel.

### 2.6 Ethical considerations and reporting guideline

Ethical approval was obtained on 24/07/2024 from the Ethical Research Committee of the University of Ilorin Teaching Hospital (ERC UITH) to collect data from the hospital; the approval reference number is UITH/CAT/189/VOL.21^B^/677. Verbal informed consent was obtained, participation was voluntary, privacy was maintained and names were not collected. The manuscript is structured in accordance with the STROBE checklist for cross-sectional studies.

## 3. RESULTS

### 3.1 Sociodemographic characteristics

A total of 180 respondents participated. Their ages ranged from 18 to 70 years, with a reported mean (standard deviation) of 37.5 (14.2) years. While all the respondents were of Yoruba ethnicity, most respondents were male (55.0%), married (65.6%), self-employed (69.4%) and had secondary education (41.7%).

### 3.2 Knowledge, attitudes and practices

Overall, 102 respondents (56.7%) were classified as having good knowledge of refuse disposal, 60 (33.3%) fair knowledge and 18 (10.0%) poor knowledge. Most of the respondents gave correct definition of refuse, 158 (87.8%); and correct definition of refuse disposal, 137 (76.1%); few however responded “I do not know” when asked what constitute proper refuse disposal. Nylons, 126 (80.2%), and solid waste 86 (54.8%) were the commonest types of waste generated. Also, the commonest source of awareness was radio, 56 (54.9%). Most of the respondents answered correctly that proper refuse disposal is important, 159 (88.3%); and that indiscriminate refuse disposal is not good, 174 (96.7%). Prevention of spread of disease, 125 (78.6%), and neat and healthy environment 114 (71.7%) were the highest responses recorded for the importance of proper refuse disposal. Most of the respondents are aware that indiscriminate refuse disposal negatively affects their environment, 153 (85.0%). Disease outbreak, 125 (78.6%), was the highest response given for the effect of indiscriminate refuse disposal on environment; while erosion recorded the least response 5 (3.3%). Most of the respondents are aware that indiscriminate refuse disposal negatively affects their health and expose them to diseases, 155 (86.1%). Cholera, 137 (88.4%), was the highest response given for the effect of indiscriminate refuse disposal on health; while fever recorded the least response 1 (0.6%). Less than half of the respondents are aware that there are laws against indiscriminate refuse disposal, 67 (37.2%). Fines, 52 (77.6%), was the most common punishment or law against indiscriminate refuse disposal that they are aware of; while “no burning” was the least 7 (10.4%). Most of the respondents are aware of methods of refuse disposal, 168 (93.3%). Open dumping, 168 (100.0%), was the commonest method known; while “drainage dumping” recorded the least response 6 (3.5%).

Eighty-seven respondents (48.3%) had a negative attitude, while 93 (51.7%) had a positive attitude. Practice was classified as good in 81 respondents (45.0%) and poor in 99 (55.0%). The most frequently reported disposal methods were incineration (50.6%), disposal into drainage channels (30.6%), recycling (27.2%) and open burning (21.7%). Multiple methods could be reported; therefore, percentages are not mutually exclusive. Only four respondents (2.2%) reported the presence of public refuse-disposal containers in their area. The leading barriers were absence of containers (37.2%) and distance to containers (28.9%); 27.8% reported no challenge.

### 3.3 Associations between knowledge, attitudes and practices

#### Hypothesis One

The level of practice of refuse disposal among the respondents has no significant influence on their level of attitude toward it.

#### Hypothesis Two

The level of knowledge on refuse disposal among the respondents has no significant influence on their level of practice

#### Hypothesis Three

The level of knowledge on refuse disposal among the respondents has no significant influence on their level of attitude towards it.

From the chi-square, it was deduced that knowledge level was associated with practice level (χ^2^=22.122; p<0.001). Knowledge level was also associated with attitude (χ^2^=8.221; p=0.016). Practice level was associated with attitude (χ^2^=5.970; p=0.015).

**Table 1.** Sociodemographic characteristics of respondents (n=180). Percentages may not sum to 100 because of rounding.

| **Characteristic** | **Category** | **n (%)** |
|---|---|---|
| Age (years) | 18–30 | 63 (35.0) |
|  | 31–40 | 50 (27.8) |
|  | 41–50 | 30 (16.7) |
|  | 51–70 | 37 (20.6) |
| Sex | Male | 99 (55.0) |
|  | Female | 81 (45.0) |
| Marital status | Married | 118 (65.6) |
|  | Single | 55 (30.6) |
|  | Divorced/separated or widowed | 7 (3.9) |
| Occupation | Self-employed | 125 (69.4) |
|  | Trading | 23 (12.8) |
|  | Other occupations | 32 (17.8) |
| Education | No formal education | 40 (22.2) |
|  | Primary | 32 (17.8) |
|  | Secondary | 75 (41.7) |
|  | Tertiary | 33 (18.3) |

Note: Sociodemographic characteristics of respondents (n=180). Percentages may not sum to 100 because of rounding. A total of 180 respondents participated. Their ages ranged from 18 to 70 years, with a reported mean (standard deviation) of 37.5 (14.2) years. Most respondents were male (55.0%), married (65.6%), self-employed (69.4%) and had secondary education (41.7%).

**Table 2.** Chi-square tests of associations among knowledge, attitude and practice (n=180).

| **Association** | **χ<sup>2</sup>** | **p-value** | **Interpretation** |
|---|---|---|---|
| Knowledge and practice | 22.122 | <0.001 | Statistically significant |
| Knowledge and attitude | 8.221 | 0.016 | Statistically significant |
| Practice and attitude | 5.970 | 0.015 | Statistically significant |

Note: Chi-square tests of associations among knowledge, attitude and practice (n=180). Knowledge level was associated with practice level (χ<sup>2</sup>=22.122; p<0.001). Knowledge level was also associated with attitude (χ<sup>2</sup>=8.221; p=0.016). Practice level was associated with attitude (χ<sup>2</sup>=5.970; p=0.015).

## 4. DISCUSSION

### 4.1 Discussion

In a 2023 study done on refuse disposal methods among residents of Emohua Local Government Area (LGA) of Rivers State, the respondents who were within the age group of 23-27 years (M=2.60, SD=1.06) did a better practice of refuse disposal methods. While the level of education (F3, 483=4.130, p<0.05) and occupation (F3, 4.573, P<0.05) were statistically significant in getting this result, age had no statistical significance in the practice of refuse disposal methods among the people of Emohua LGA (F3,483=2,420, p> 0.05).^[6]^ The mean plot for the riverain Emohua LGA showed that residents who had a primary level of education did more practice of refuse disposal (M=2.610). Also, the results of the study revealed that these residents with a primary level of education indicated that they dump their refuse into the river, gutter or drains.^[6]^

Moving up geographically to Ilorin, Ilorin experiences a tropical continental climate with distinct wet and dry seasons. The wet season spans from March to October. Neglecting waste disposal leads to severe air pollution and foul odors, especially during the rainy season. In 2024, a study on environmental protection policies for sustainable waste management was conducted was in Kwara and Oyo States by Amin Amin et al. It was discovered that there is lack of effective strategies for solid wastes disposal in both states studied.^[2]^ As for this current study, it was observed that slightly more than half of respondents had good knowledge of refuse disposal, yet more than half were classified as having poor disposal practices. This difference suggests that awareness alone may not be sufficient to achieve appropriate household waste management. Availability, convenience and reliability of collection services are likely to be important contextual factors; in this study, almost all respondents reported that public refuse containers were absent from their area.^[4,5,6]^

Negative attitudes were reported by 48.3% of respondents. Although the proportion with a positive attitude was slightly higher, the coexistence of negative attitudes and poor practices indicates that behavioural and structural barriers may operate together. The reported use of incineration, disposal into drainage channels and open burning is particularly important for local environmental-health planning.

The most frequently reported barriers were lack of waste containers and distance to available containers. These findings point to a service-access problem as well as a need for public education. Recommendations focused solely on changing individual behaviour may have limited impact if residents lack accessible collection points or regular collection services. Local authorities and waste-management providers should consider the placement and maintenance of communal containers, predictable collection schedules and accessible information about approved disposal options.^[3,5,6]^

From the chi-square analyses, there are statistically significant associations between knowledge and practice, knowledge and attitude, and practice and attitude. These findings support the relevance of the three domains to one another.

### 4.2 Strengths and limitations

The study provides community-specific data from 180 adult residents and assessed knowledge, attitudes, reported practices and perceived barriers within one questionnaire. Limitations include the cross-sectional design, which precludes causal inference (that is, it cannot establish temporal sequence or causation); reliance on self-reported behaviour, which may be affected by recall and social-desirability bias; and restriction to one community, which limits generalisability. In addition, the reported p-values do not quantify the magnitude of the associations between variables. Future analyses could address that and also report effect sizes and confidence intervals and examine potential confounding by education, age, occupation and access to waste services.

## 5. CONCLUSION AND IMPLICATIONS FOR TRANSLATION

Among adults in Okelele community, good knowledge of refuse disposal was more common than good disposal practice. Poor practices were frequently reported, and the main barriers were the absence of public waste containers and the distance to available containers. Knowledge, attitude and practice levels were statistically associated, although causality cannot be inferred. Public-health action should employ effective strategies and combine community education with accessible waste-collection infrastructure, reliable collection schedules and consistent implementation of local environmental sanitation policies and regulations.

## Data Availability

All data produced in the present study are available upon reasonable request to the authors.

https://docs.google.com/document/d/1IPd02OL8aFvz8h5LI2J0vYNnsWr_5Kwr/edit?usp=sharing&ouid=115863318443348500928&rtpof=true&sd=true

https://docs.google.com/document/d/1_MkWG5ZWKrDtfwsVjdCyOUHLLRvPzbpa/edit?usp=sharing&ouid=115863318443348500928&rtpof=true&sd=true

## ACKNOWLEDGMENTS

The authors acknowledge the residents of Okelele community who participated in the study and the Department of Epidemiology and Community Health, University of Ilorin.

## COMPLIANCE WITH ETHICAL STANDARDS

Conflicts of interest: The authors declare no competing interests. Financial disclosure: Nothing to declare.

### Funding/support

There was no funding for this study.

### Ethics approval

Ethical approval was obtained on 24/07/2024 from the Ethical Research Committee of the University of Ilorin Teaching Hospital (ERC UITH); the reference number is UITH/CAT/189/VOL.21^B^/677.

### Declaration of patient/participant consent

Verbal informed consent was obtained from all participants before data collection. Participation was voluntary, and confidentiality was maintained.

Use of artificial intelligence (AI)-assisted technology for manuscript preparation: AI-assisted technology was used to reorganise and edit the manuscript into the journal’s requested structure. The authors however take full responsibility for all content of the article.

### Disclaimer

None.

## KEY MESSAGES

- Good knowledge of refuse disposal was reported by 56.7% of respondents, but 55.0% had poor disposal practices.
- Lack of public waste containers and distance to available containers were the leading reported barriers.
- Knowledge, attitude and practice were significantly associated, supporting combined education and service-access interventions.

## TABLES AND FIGURES

**Figure 1.**
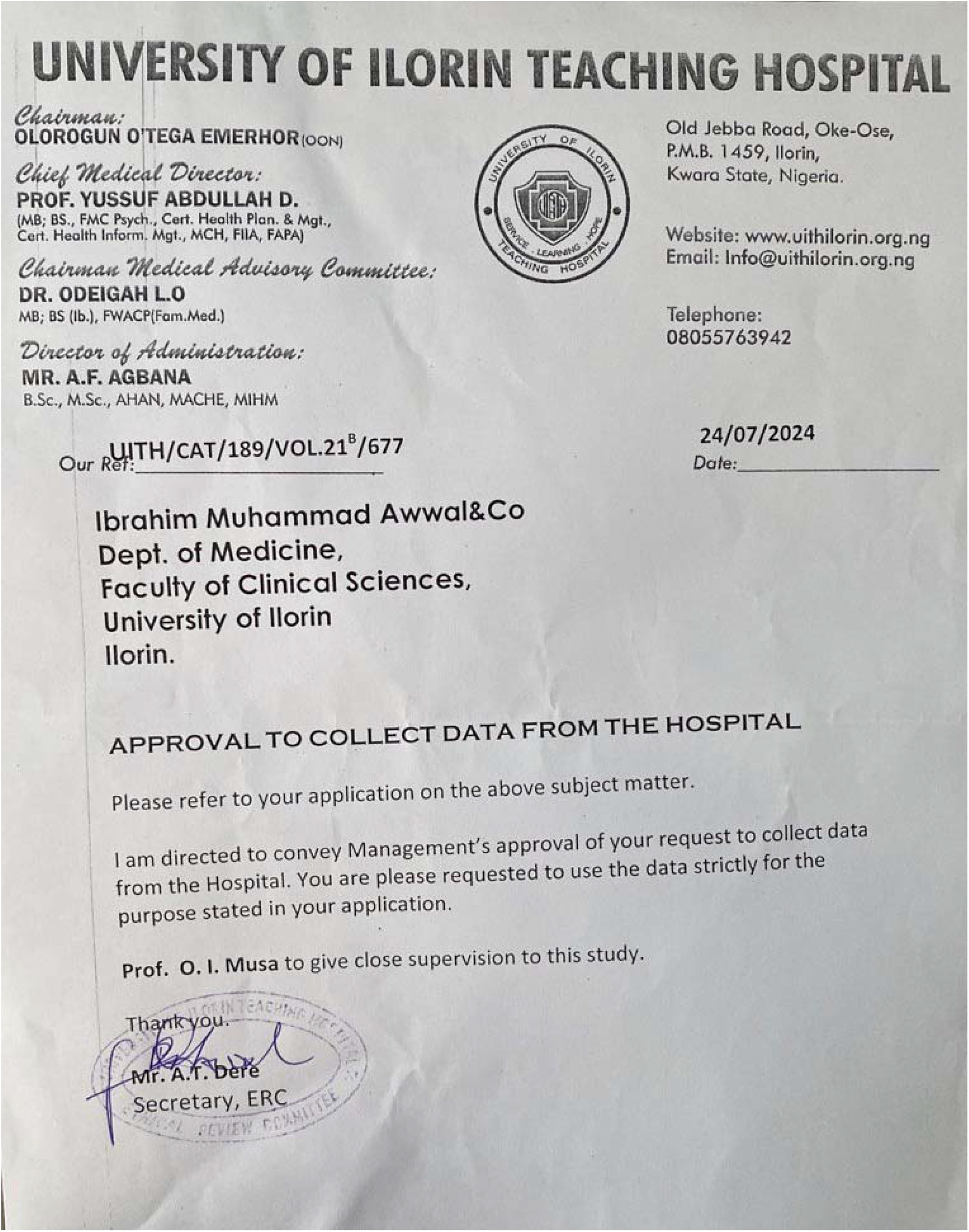
Ethical Approval from ERC UITH. Ethical approval was obtained on 24/07/2024 from the Ethical Research Committee of the University of Ilorin Teaching Hospital (ERC UITH); the reference number is UITH/CAT/189/VOL.21^B^/677.

